# Integration of lung tissue proteomics and genome-wide association data to identify lung cancer susceptibility proteins and potential drug targets

**DOI:** 10.64898/2026.06.18.26355973

**Authors:** Shuai Xu, Jiajun Shi, Xiao-Ou Shu, Ran Tao, Yongchao Dou, Xingyi Guo, Wanqing Wen, Yaohua Yang, Bing Zhang, Jie Wu, Stephen A. Deppen, Bingshan Li, Wei Zheng, Jirong Long, Qiuyin Cai

**Author notes:** Contributed equally (Shuai Xu, Jiajun Shi). **Corresponding authors contact information:** Qiuyin Cai, M.D., Ph.D. Department of Medicine, Vanderbilt Epidemiology Center Vanderbilt-Ingram Cancer Center, Vanderbilt University School of Medicine; Jirong Long, Ph.D., Department of Medicine, Vanderbilt Epidemiology Center Vanderbilt-Ingram Cancer Center Vanderbilt University School of Medicine.

## Abstract

**Background:** Proteins directly impact disease development and act as drug targets. Therefore, we integrated genomic and lung tissue proteomics data to identify lung cancer susceptibility proteins, elucidating genetic mechanisms and candidate drug targets.

**Method:** We profiled the proteome and genome in non-neoplastic lung tissue from 200 lung cancer patients. Using this data, we constructed genetic models to predict abundance across the proteome in lung tissue. We applied these models to genome-wide association study (GWAS) data from 55,174 lung cancer cases and 1,294,174 controls to evaluate their associations with the risk of lung cancer, overall and by major histological subtypes. Bayesian colocalization and Mendelian randomization (MR) analyses were used to prioritize putative causal proteins, which were cross-referenced with three main drug-protein databases to identify potential therapeutic targets.

**Results:** We identified 29 proteins associated with lung cancer risk at a false discovery rate < 5%, including 25 for overall lung cancer, two (AQP3 and IL18) specifically for adenocarcinoma, and another two (HMGN2 and HLA-DMB) for squamous cell carcinoma. Of them, genes encoding 17 proteins reside at least 2Mb away from any known GWAS risk loci, including 14 for overall lung cancer (HYI, GPX1, GMPPB, DSP, HDDC2, MTCH2, SUOX, JMJD7, PDIA3, IL16, IQGAP1, SULT1A2, ARHGAP27, and TYMP) and three for subtypes (AQP3, IL18, and HMGN2). Among the 12 proteins located within the known risk loci, EPHX2, CLDN18, PSMD5, and CYP2S1 proteins showed an association independent of the proximal GWAS-identified lead variant. Colocalization and/or MR analysis suggested 11 potential causal proteins. Five of these candidate causal proteins (DSP, CLDN18, IQGAP1, IL18 and TYMP) are targeted by nine drugs already approved by the FDA or in phase III trials.

**Conclusion:** Our study identified novel lung cancer susceptibility proteins and potential drug targets, offering valuable insights into lung cancer biology and future translational utilities.

## Introduction

Lung cancer is the second most common cancer and the leading cause of cancer mortality among both men and women globally (1,2). While cigarette smoking is the primary risk factor, genetic susceptibility plays a critical role in lung cancer development for both smokers and non-smokers. A twin study estimated that genetic factors account for roughly 20% of susceptibility (3). Although genome-wide association studies (GWASs) have identified approximately 100 risk loci for lung cancer (4,5), most of these loci lie in noncoding regions, making it difficult to identify the underlying causal genes.

Transcriptome-wide association studies (TWASs) were developed to bridge this gap by linking genetically regulated gene expression to lung cancer risk and have identified more than 100 putative susceptibility genes (6–8). However, candidate genes have not yet been discovered for several GWAS loci associated with lung cancer risk. Incorporating previously unexplored omics data could reveal additional insights into genetic mechanisms in GWAS loci.

Proteins directly impact biological and cellular functions and ultimately determine phenotypes rather than gene transcripts. Although gene expression levels were commonly used as proxies for protein abundance, it is well recognized that gene expression levels are often weakly correlated with protein abundance, and distinct genetic regulations have been observed between transcription and translation in most human tissue (9,10). This discordance underscores the importance of investigating potential lung cancer pathogenesis mechanisms at the protein-level. Proteome-wide associations (PWAS) extend the TWAS computational framework by assessing associations between genetically predicted protein abundance and disease risk. We previously used this approach and identified 10 plasma protein biomarkers in relation to the risk of overall lung cancer or adenocarcinoma (11). However, plasma proteins may not capture the tissue-specific etiological mechanisms of this malignancy. For example, many proteins expressed in lung tissue may not be detectable in plasma samples, and the correlations between plasma and lung tissue are modest. Therefore, it is crucial to utilize lung-tissue proteome data to identify lung cancer risk proteins. The findings are crucial to advance our understanding of lung cancer etiology and inform precision prevention strategies.

Beyond offering novel etiological insights into lung carcinogenesis, identifying proteins in lung tissue has direct therapeutic implications. As most drugs usually modulate the activities of their target proteins, identifying novel proteins has the potential of accelerating therapeutic development (12). Here, we presented the findings from the first large-scale lung tissue-based PWAS to identify candidate lung cancer susceptibility proteins as potential therapeutic targets for lung cancer prevention and treatment.

## Method

### Proteomics data profiling and quantification

We profiled the proteome of fresh-frozen treatment-naïve tumor-adjacent normal lung tissue from 200 lung cancer patients using next-generation label-free Data Independent Acquisition (DIA) mass spectrometry (MS). In brief, the main experimental pipeline includes protein extraction, enzymatic digestion, and high-pH reversed-phase fractionation. First, we constructed a comprehensive spectral library using Data Dependent Acquisition (DDA) to capture non-redundant, high-quality peptide signatures from samples. Subsequently, individual samples were analyzed via nanoscale liquid chromatography coupled to tandem MS on a Q-Exactive HFX system operating in DIA mode.

The DDA raw data was processed using the Andromeda search engine within MaxQuant (13) against reviewed human protein sequences from UniProt database (14), and the results were used to generate the comprehensive spectral library required for the subsequent DIA analysis. Only peptides satisfying a false discovery rate (FDR) < 1% were retained to construct the final spectral library. We employed the mProphet algorithm (15) to conduct rigorous analytical quality control, ensuring robust and reliable quantification. DIA data were analyzed using iRT peptides for precise retention time calibration.

### Proteomics data quality control and processing

We first excluded proteins with missing data among at least 50% of samples and did not perform any imputation, as the Clinical Proteomic Tumor Analysis Consortium (CPTAC) recommended (16).

Only proteins with encoding genes located on autosomes were included in the downstream analysis. Participants with a missing rate > 40% were further excluded. We then performed log_2_ transformation on protein abundance data. Uniprot ID was mapped to Ensembl ID and then chromosome and position information of each corresponding protein-coding gene was acquired using the R *biomaRt* Bioconductor package. We next performed rank-based inverse normalization on log2-transformed protein abundance data.

### Genotype data quality control

Lung tissue samples were genotyped using the Multi-Ethnic Genotyping Array at Vanderbilt University Medical Center. The genotype data quality control, processing, and imputation procedures were described in detail in our previous work (6). Briefly, we excluded genetic variants with a minor allele frequency (MAF) < 1%, a Hardy-Weinberg equilibrium *P* < 10^-6^, a missing rate > 5%, and a consistency rate of < 98% among duplicate samples. We removed individuals with a missing rate > 5% and relatedness (pi_hat>0.125). Then genotype data were imputed using the Trans-Omics for Precision Medicine (TOPMed) version 2.3 as the reference panel. After imputation, genetic variants with high imputation quality scores (R^2^ > 0.8), MAF > 5%, and availability in lung cancer risk GWAS summary statistics were retained. Allele frequencies of palindromic variants were compared with those of European ancestry individuals from the 1000 Genome Project. No palindromic variants with huge (> 0.2) differences in allele frequencies were identified.

### Building genetic prediction models

We followed our previously established protocol to build protein prediction models (6,11,17). To account for the potential confounding effects, residuals were derived by regressing normalized protein abundance on age (continuous), biological sex (male or female), smoking status (ever or never), top three genetic principal components (continuous), and 30 probabilistic estimation of expression residuals (PEER) factors (continuous) (18). We further conducted rank-based inverse normalization on them to ensure the residuals were normally distributed.

Genetic variants located within 500 kilobases upstream and downstream of each gene encoding the corresponding protein were selected. The elastic net models (*α* = 0.5) with nested cross-validation were constructed to predict the abundance level of each protein implemented in the R *glmnet* package. First, we estimated the minimum cross-validated errors by five-fold cross-validation in the inner loops. Then we applied the minimum errors and computed the Spearman correlation between the observed and the predicted abundance to evaluate prediction performance in the outer loops. Reliable prediction models were defined as Spearman’s rho > 0.1 and with significance at *P* < 0.05 (6).

### Statistical analysis for genetically predicted protein levels with lung cancer risk

For proteins with reliable prediction models, each reliable model was applied to the lung cancer GWAS summary statistics using the S-PrediXcan framework (19) to evaluate the associations of genetically predicted protein abundance levels with lung cancer risk. The formula is shown below:

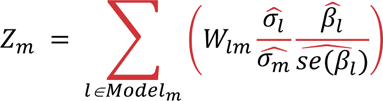

The *Z* score estimates the direction and strength of associations between the genetically predicted protein abundance level and lung cancer risk. W_lm_ represents the weight of genetic variant *l* in the prediction model for protein *m*. Terms 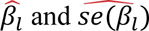 refer to the regression coefficient and the corresponding standard error for lung cancer risk associated with genetic variant *l* in the GWAS meta-analysis summary statistics, respectively. 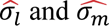 denote the estimated variances of genetic variant *l* and the predicted abundance level of protein *m*, respectively. We also computed the correlations between genetic variants included in the prediction models. Significance level was defined as false discovery rate (FDR) < 5% to account for the multiple testing burdens.

### Conditional analysis of GWAS index variants

We further evaluated whether associations between the genetically predicted abundance of identified proteins and lung cancer risk were independent of the nearby GWAS-identified lung cancer lead variants. First, we performed conditional and joint analysis (COJO) on lung cancer risk GWAS summary statistics, adjusting for the proximal GWAS-identified index variants (20). COJO analysis also leverages the linkage disequilibrium (LD) reference among the European samples from the 1000 Genomes Project. The lung cancer GWAS-identified index variants were acquired from the recent large-scale GWASs of lung cancer risk (5,21–25). We then applied those prediction models to the adjusted GWAS summary statistics using the S-PrediXcan algorithm. The independent association was defined as Bonferroni-corrected *P* < 0.05 after conditional analysis.

### Protein-set enrichment analysis

For the proteins associated with lung cancer risk, we constructed a protein-protein interaction (PPI) network to investigate the connectiveness of these proteins using through STRING PPI database (26). To identify high-confidence interactions, we set a minimum interaction score of 0.7. Further we performed protein-set enrichment analysis with respect to Gene Ontology (GO) annotations and *Reactome* pathways. GO terms or pathways with more than two proteins and exhibiting an FDR < 5% were considered to be significantly enriched.

### Lung alveolar cell type specificity

We evaluated lung alveolar cell type-specific protein abundance using Deep Visual Proteomics data from the Human Protein Atlas (HPA) (27). PWAS-identified proteins were classified into four categories based on the HPA definitions: (1) alveolar cell type enriched (≥ fourfold higher abundance in alveolar cell type compared to all other cell types), (2) group enriched (enriched in a group of the alveolar cell type and up to nine other cell types), (3) alveolar cell type enhanced (≥ fourfold higher abundance in the alveolar cell type compared to the mean of all other cell types), and (4) not detected in alveolar cell type. Proteins meeting criteria 1 or 2 were all classified as alveolar cell type enriched.

### Comparison of TWAS with PWAS

We first acquired the associations results between genetically predicted gene expression levels of those significant proteins and lung cancer risk from our recent TWAS study among 314 participants of European Ancestry from the same study population (6). We compared the results of TWAS and PWAS to assess whether transcriptomic- and proteomic-level associations were consistent. We then obtained gene expression level (Transcript Per Million) data from these 200 participants with available DIA proteomics data. We calculated the Spearman’s rho between protein abundance and its protein-coding gene expression level. Bonferroni corrected *P* < 0.05 was considered as the significance level. Finally, to explore whether protein expression levels mediate the associations between gene expression and cancer risk, we jointly analyzed results from the above TWAS and PWAS associations and correlations.

Specifically, a coherent gene-protein-cancer relationship requires the directions of protein-cancer, gene-protein, and gene-cancer associations to be reasonably consistent.

### *In silico* causal inference

For the identified proteins, we first conducted colocalization analyses to determine whether protein abundance and lung cancer risk shared a causal genetic variant. We performed c*is*-pQTL analysis using QTLtools (28), adjusting for the above-mentioned confounders. This analysis was conducted using the R *coloc* package under a single causal variant assumption.(29) We used the default prior probabilities (PP) of P_1_ = P_2_ = 10^−4^ and P_12_ = 10^−5^. A posterior probability (PP) of hypothesis 4 (causal variants shared by both traits) > 0.8 (80%) was defined as evidence for sharing a genetic signal (30).

Summary-data-based Mendelian randomization (SMR) was then used to identify potential causal associations of protein abundance with lung cancer risk (31). We set *P* of < 1×10^-5^ to select the top *cis*-pQTLs and used other default parameters (30). We also conducted the heterogeneity in dependent instruments (HEIDI) test to evaluate the absence of linkage. The proteins with Bonferroni-corrected *P_SMR_* < 0.05 and HEIDI *P* > 0.05 were considered potential causal protein markers for lung cancer risk.

### Querying drug targets

To identify potential therapeutic targets, we evaluated candidate causal proteins using three major drug-protein databases, including DrugBank (version 5.1.17) (32), ChEMBL (version 36, updated on July 28, 2025) (33), and TTD (34). From these databases, we extracted drugs and their annotated protein targets and linked them to our identified potential causal proteins. Our mapping focused mainly on drugs approved or under Phase II or III clinical trials.

## Results

### Protein prediction model building

The analytical workflow and design of this study is illustrated in Figure 1. We generated and used DIA proteomics data from pathologically confirmed normal lung tissues of 200 European Americans to construct genetic prediction models for protein abundance with matched genetic data. After quality control, 5,583 autosomal proteins with a missing rate less than 50% across 200 participants were retained for downstream prediction model building. The participants had a median missing rate in protein abundance of 6.2%, with an interquartile [IQR] of 3.6% to 13.5%. Of the 5,583 autosomal proteins, genetic prediction models were built for 2,751 proteins. Among them, 805 proteins were predicted reliably (Spearman’s rho > 0.1 and *P* < 0.05), representing the largest lung-tissue proteomic prediction models to date. Reliable prediction models were built on a median of 22 genetic variants (IQR: 12–37) and a median of 200 participants (IQR: 195–200).

**Figure 1.**
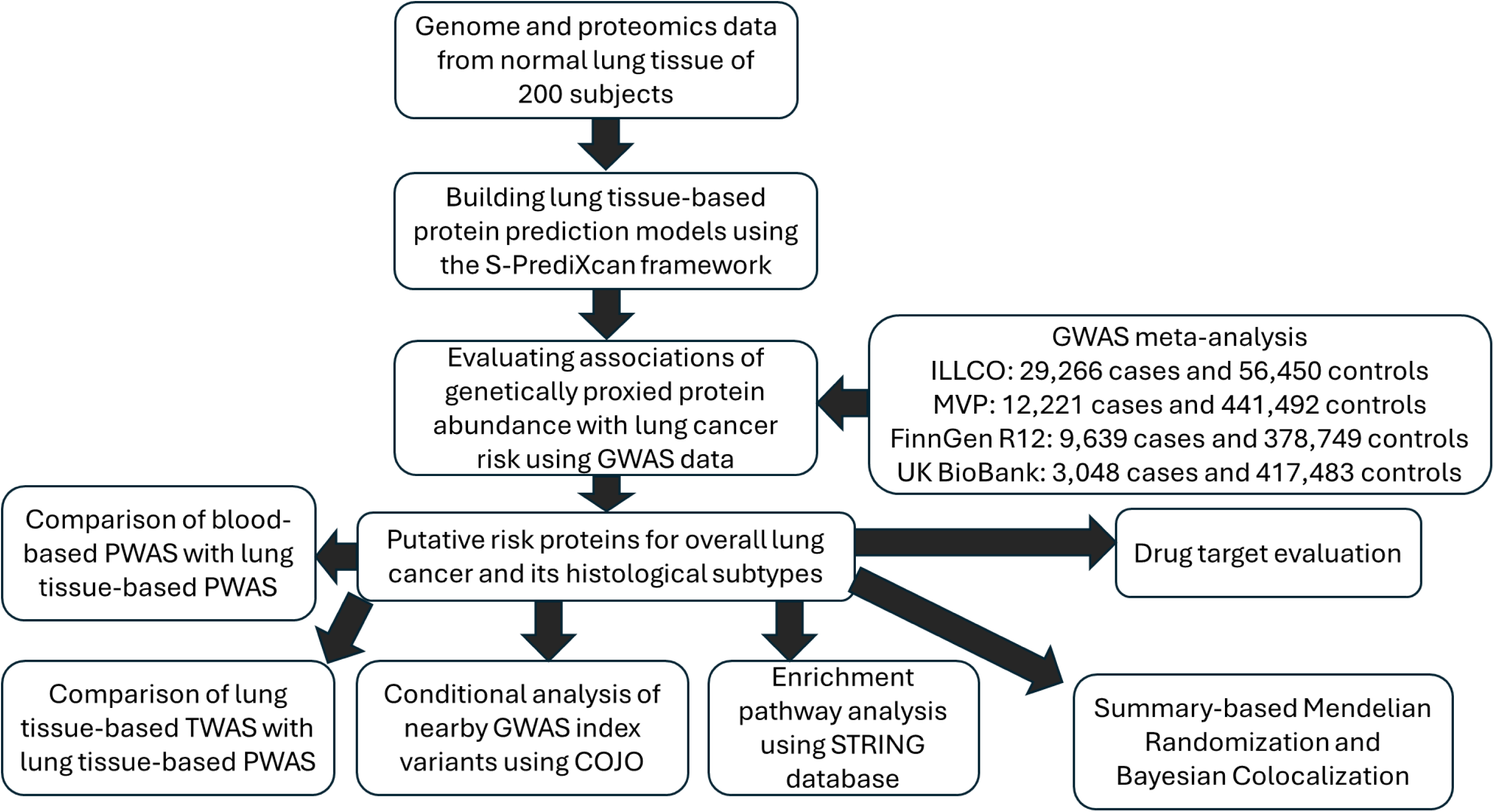
Analytic schematic and flowchart.

### Predicted Proteins associated with overall lung cancer risk

By applying 805 reliable prediction models to the GWAS summary statistics, we identified 25 proteins whose genetically predicted abundance was associated with overall lung cancer risk at FDR < 5%. Among them, encoding genes for 14 identified proteins were located at least 2 Mb away from GWAS-identified genetic variants for lung cancer risk, indicating that these proteins were located in potential new risk loci. These 14 proteins are HYI, GPX1, GMPPB, DSP, HDDC2, MTCH2, SUOX, JMJD7, PDIA3, IL16, IQGAP1, SULT1A2, ARHGAP27, and TYMP (Table 1). Of them, 11 proteins (HYI, GPX1, HDDC2, MTCH2, SUOX, JMJD7, IL16, IQGAP1, SULT1A2, ARHGAP27, and TYMP) have not been reported to be associated with lung cancer risk previously in either lung tissue-based TWAS or plasma PWAS (6,8,11,35).

**Table 1.** Fourteen proteins associated with overall lung cancer risk located at least 2Mb away from any known lung cancer GWAS-identified risk loci.

| Protein | Cytoband | Z | P |
| --- | --- | --- | --- |
| HYI <sup>a</sup> | 1p34.2 | 3.94 | 8.00×10<sup>-5</sup> |
| GPX1 <sup>a</sup> | 3p21.31 | 3.75 | 1.76×10<sup>-4</sup> |
| GMPPB | 3p21.31 | 4.81 | 1.55×10<sup>-6</sup> |
| DSP | 6p24.3 | -4.23 | 2.37×10<sup>-5</sup> |
| HDDC2 <sup>a</sup> | 6q22.31 | -3.51 | 4.45×10<sup>-4</sup> |
| MTCH2 <sup>a</sup> | 11p11.2 | 4.09 | 4.30×10<sup>-5</sup> |
| SUOX <sup>a</sup> | 12q13.2 | -3.63 | 2.85×10<sup>-4</sup> |
| JMJD7 <sup>a</sup> | 15q15.1 | 4.28 | 1.88×10<sup>-5</sup> |
| PDIA3 | 15q15.3 | 3.93 | 8.60×10<sup>-5</sup> |
| IL16 <sup>a</sup> | 15q25.1 | 3.83 | 1.28×10<sup>-4</sup> |
| IQGAP1 <sup>a</sup> | 15q26.1 | -3.36 | 7.67×10<sup>-4</sup> |
| SULT1A2 <sup>a</sup> | 16p11.2 | -4.63 | 3.63×10<sup>-6</sup> |
| ARHGAP27 <sup>a</sup> | 17q21.31 | 3.34 | 8.52×10<sup>-4</sup> |
| TYMP <sup>a</sup> | 22q13.33 | -3.62 | 2.97×10<sup>-4</sup> |

Kb = Kilobases; GWAS = Genome-wide association study.
<sup>a</sup>Proteins that have not been reported in any previous lung-tissue-based TWAS or blood-based PWAS of lung cancer.

The genes encoding the remaining 11 identified proteins are located within 2Mb of the GWAS-identified lung cancer index variants (Table 2). Conditional analysis revealed that four of these 11 proteins remained significant associations with lung cancer risk after adjustment for GWAS–identified lung cancer risk variants, suggesting that their associations are independent of known GWAS signals. These four proteins are EPHX2 (*P* = 2.54×10^−6^and *P*_conditional_ = 9.02×10^−6^), CLDN18 (*P* = 6.62×10^−5^ and *P*_conditional_ = 6.07×10^−5^), PSMD5 (*P* = 2.30×10^−4^and *P*_conditional_ = 1.61×10^−4^), and CYP2S1 (*P* = 2.00×10^−7^ and *P*_conditional_ = 1.08×10^−3^) (Table 2). Among these four proteins, EPHX2, PSMD5, and CYP2S1 have not been reported to be associated with lung cancer risk previously in either lung tissue-based TWAS or plasma PWAS. Of the remaining seven proteins whose associations were driven by the GWAS-identified lung cancer index variants, HLA-DMA and C4B have not been reported in either lung tissue-based TWAS or plasma PWAS for lung cancer risk.

**Table 2.** Eleven proteins associated with overall lung cancer risk located within the known lung cancer GWAS risk loci.

| Protein | Cytoband | Z | *P* | *P*<sub>adjust</sub> |
| --- | --- | --- | --- | --- |
| CASP8 | 2q33.1 | -4.87 | $1.10 \times 10^{-6}$ | 0.46 |
| CLDN18 | 3q22.3 | -3.99 | $6.62 \times 10^{-5}$ | $6.07 \times 10^{-5}$ |
| HLA-A | 6p22.1 | -4.09 | $4.24 \times 10^{-5}$ | 0.34 |
| FLOT1 | 6p21.33 | -6.41 | $1.48 \times 10^{-10}$ | $3.80 \times 10^{-3}$ |
| C4A | 6p21.33 | -5.45 | $5.13 \times 10^{-8}$ | 0.42 |
| C4B <sup>a</sup> | 6p21.33 | -3.95 | $7.84 \times 10^{-5}$ | 0.04 |
| HLA-DMA <sup>a</sup> | 6p21.32 | -4.29 | $1.81 \times 10^{-5}$ | 0.60 |
| EPHX2 <sup>a</sup> | 8p21 | -4.70 | $2.54 \times 10^{-6}$ | $9.02 \times 10^{-6}$ |
| PSMD5 <sup>a</sup> | 9q33.2 | -3.68 | $2.30 \times 10^{-4}$ | $1.61 \times 10^{-4}$ |
| CTSH | 15q25.1 | 4.50 | $6.88 \times 10^{-6}$ | 0.86 |
| CYP2S1 <sup>a</sup> | 19q13.2 | -5.20 | $2.00 \times 10^{-7}$ | $1.08 \times 10^{-3}$ |

*P*<sub>adjust</sub>: *P* values after adjusting for the nearby GWAS index variants.
<sup>a</sup> Proteins that have not been reported in any previous lung-tissue-based TWAS or blood-based PWAS of lung cancer.

In total, 16 identified proteins have not been reported to be associated with lung cancer risk previously. Additionally, nine proteins (CASP8, GMPPB, DSP, PDIA3, C4A, FLOT1, CTSH, HLA-A, and CLDN18) have been identified in previous TWASs of lung cancer risk. These support the validity of our results while providing complementary protein-level evidence for lung cancer susceptibility.

### Associations by lung cancer histological subtypes

Different histological subtypes may have distinct genetic mechanisms. Therefore, we performed stratification analysis to identify proteins specifically associated with the risks of three major histological subtypes, namely adenocarcinoma (LUAD), squamous cell carcinoma (LUSC), and small cell carcinoma (SCLC). The AQP3 and IL18 proteins were solely significantly associated with LUAD risk, while the HMGN2 and HLA-DMB proteins were only significantly associated with LUSC risk at FDR < 5% (Table 3). Of these four histological subtype-specific proteins, two LUAD-associated (IL18 and AQP3) and one LUSC-associated (HMGN2) risk proteins were located more than 2Mb from the known lung cancer GWAS index variants, nominating the potential novel risk loci. Of note, one LUAD-(IL18) and all LUSC-(HMGN2 and HLA-DMB) specific risk proteins were not reported in any previous lung-tissue-based TWAS or blood-based PWAS of lung cancer. However, we did not find any protein significantly associated with SCLC risk, likely due to limited statistical power from smaller GWAS sample sizes.

**Table 3.**
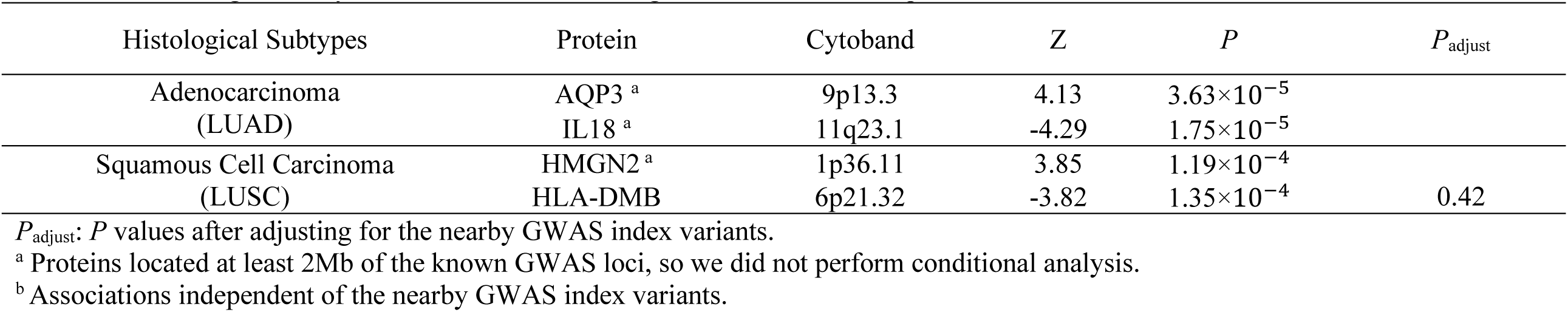
Proteins significantly associated with risk of lung adenocarcinoma or squamous cell carcinoma.

We next evaluated heterogeneity in association of the 25 overall lung cancer risk proteins across three major histological subtypes. Directions of these associations were consistent across all subtypes. At FDR < 5%, five proteins were associated with LUAD (C4A, CASP8, CTSH, ARHGAP27, and TYMP) and six with LUSC (HYI, HLA-A, FLOT1, C4A, C4B, and HLA-DMA) (Table 4). Additionally, 11 proteins for LUAD risk, seven for LUSC risk, and six for SCLC risk showed a significant association at nominal *P* < 0.05 but not FDR < 5%.

**Table 4.**
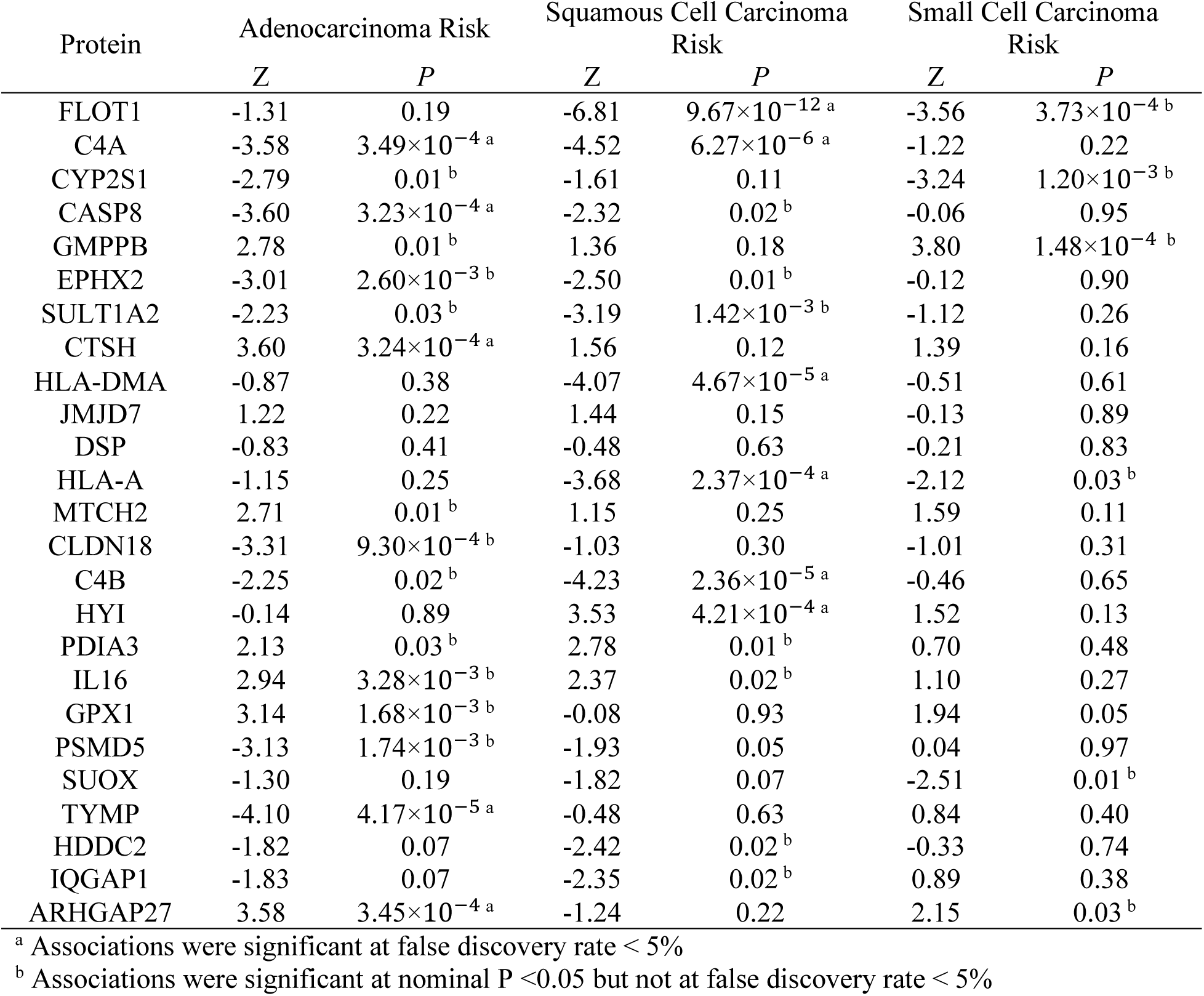
Associations between the risk of three major lung cancer histological subtypes and 25 identified proteins mentioned in Tables 1 and 2.

### Functional characterization and Cell type specificity

We interrogated the STRING PPI database to characterize functional connections and enrichment among 29 PWAS-identified proteins. Among them, we found two high-confidence protein communities based on PPI (Figure S1). Overall lung cancer-associated risk proteins were significantly enriched across one Gene Ontology (GO) cellular component, and three *Reactome* pathways at FDR < 5% (Table S1a). In addition, LUSC risk proteins were enriched in 17 GO biological processes, four GO cellular components, and a *Reactome* pathway at FDR < 5% (Table S1b). Most are immune-related biological processes and pathways. For example, *classical-complement-pathway C3/C5 convertase complex* and *activation of C3 and C5* were identified and related to the complement pathway. Notably, complement pathway inhibition has been shown to suppress lung tumor growth *in vivo* (36).

We additionally found six proteins were detectable in alveolar cell types. Among these, three proteins (CLDN18, CYP2S1, and CTSH) were enriched in alveolar cell types and three proteins (C4A, C4B, and HLA-DMA) were classed as alveolar cell type enhanced. Of note, alveolar type I and II are well-known cells of origin for lung adenocarcinoma.

### Comparison of TWAS with PWAS

To evaluate whether PWAS consistently reflected transcriptomic-level associations, we compared our findings with TWAS results derived from the same GWAS summary statistics and population.

Overall, a moderate correlation (Spearman’s rho=0.58, *P* <0.001) in effect sizes between TWAS and PWAS was observed for 501 protein-coding genes that were reliably predicted at both transcriptome and proteome levels. Then we investigated TWAS results for gene coding the 29 PWAS-identified proteins, so we set the significance threshold as *P* < 1.72×10^−3^ (0.05/29) to account for multiple testing. Among 25 proteins associated with overall lung cancer risk, 13 protein-coding genes showed significant TWAS results with concordant effect directions of PWAS at *P* < 1.72×10^−3^ (Table S2). In contrast, C4B exhibited significantly opposite association directions between transcriptomic and proteomic levels. For two LUAD-specific proteins, both AQP3 and IL18 proteins showed significant transcriptome-level associations with the same direction of effect as PWAS.

Integrating transcriptome and proteomics data, we identified nine gene–protein–cancer trios with consistent TWAS and PWAS association directions and significant positive protein-gene correlations for overall lung cancer (GMPPB, CLDN18, DSP, HDDC2, PSMD5, MTCH2, JMJD7, CASP8, and TYMP) and two for LUAD risk (AQP3 and IL18) (Tables S2). For example, HDDC2—negatively associated with overall lung cancer risk in PWAS—also showed an inverse TWAS association with overall lung cancer risk (Z_TWAS_ = −3.75), and its mRNA and protein levels were positively correlated (Spearman’s rho = 0.35), illustrating a gene-protein-cancer trio.

### Prioritizing potential causal proteins and drug target query

To pinpoint likely causal proteins for lung cancer risk, we applied two *in silico* analyses: colocalization and SMR. Due to the complex LD issues in the Human Leukocyte Antigen (HLA) genomic region, we excluded 6 proteins with coding genes located in HLA region from these analyses and tested the remaining 23 proteins. Of them, six were supported as the high-confidence causal proteins by both colocalization (PP.H4 > 0.8) and SMR (*P_SMR_* < 1.72×10^−3^ and *P*_HEIDI_ > 0.05), including four (MTCH2, SUOX, JMJD7, and CLDN18) for overall lung cancer and two (IL18 and TYMP) for LUAD risk (Table S3). Five additional proteins (HDDC2, DSP, HYI, IQGAP1, and PSMD5) showed suggestive evidence for potential causal associations with the risk of overall lung cancer or LUSC by a single method.

Among these 11 proteins with additional supporting evidence of the potential causal relationship for lung cancer risk, we queried three main drug-protein databases to investigate whether they were potential targets for drugs approved by the FDA or under phase II/III clinical trial. DSP, CLDN18, IQGAP1, IL18, and TYMP proteins are targets of nine drugs that have been approved or are being investigated in phase III trials (Table S4a, S4b, and S4c). These nine drugs or compounds are Artenimol, Zinc, Zinc acetate, Tipiracil, Uridine, Zolbetuximab, MAS825, Tadekinig alfa, and Uridine.

## Discussion

In this first lung tissue-based PWAS, we identified 29 novel risk proteins, including 25 proteins for overall lung cancer, two specifically for LUAD, and two specifically for LUSC. Of these 29 proteins, 17 were located in regions not reported in any previous studies, and four were associated with lung cancer risk independently from the nearby known GWAS index variants. Nine unique coherent gene-protein-cancer relationships for overall lung cancer and two coherent relationships for LUAD were identified.

Seven proteins demonstrated strong evidence of causality, and four additional proteins demonstrated suggestive evidence of causality. Of them, five potential causal proteins are linked to nine drugs that have been approved or are under phase III clinical trials. Our study identified putative susceptibility proteins and candidate drug targets, which inform the genetic mechanisms of lung carcinogenesis and potentially repurposed drugs for lung cancer prevention and treatment.

By leveraging proteomics profiling and genetic data from a large population, we identified novel lung cancer risk proteins, many of which have not been implicated by TWAS at the transcriptomic level. The roles of these proteins in lung cancer were supported by previous studies. A short hairpin RNA knockdown experiment conducted in primary human non-small cell lung cancer cells found *MTCH2* silencing decreased cell viability, proliferation, and migration, and impaired mitochondrial functions, supporting our PWAS results (37). IQGAP1 is involved in many oncogenic signaling pathways, including MAPK, Wnt, PI3K, and TGF-beta signaling through binding to different proteins (38). SYT15B-IQGAP1 interaction activates the MAPK signaling pathway to promote A549 and H1299 cell line proliferation, migration, and invasion (39). Protein expression levels of IQGAP1 in lung tumor tissue was lower compared to those in adjacent normal lung tissues in CPTAC (16), which aligns with our finding of an inverse association between expression levels of IQGAP1 and lung cancer risk. In addition, differential protein abundance analysis in CPTAC suggested that lower SUOX, SULT1A2, CYP2S1, and PSMD5 abundance was observed among lung tumor tissue compared to tumor-adjacent normal lung tissue (16).

MR analysis showed an inverse association between genetically predicted plasma levels of IL18 and the risk of overall lung cancer and LUAD (40). IL-18 plays a role in the production of IFN-γ, which can promote tumor cell apoptosis (41,42). Both gene and protein expression levels of *EPHX2* were found to be downregulated in both LUAD and LUSC (16,43). EPHX2 can promote epoxyeicosatrienoic acids to convert into dihydroxyeicosatrienoic acids, which have pro-inflammatory properties (44,45). Chronic inflammation is an essential risk factor for lung cancer. An inverse association of plasma CASP8 protein levels with lung cancer risk has been reported previously in a case-control study (46), while we observed a positive association in lung tissue. This might be attributable to the different study design, time window, and source of protein involved. Further research is warranted to further investigate the underlying biological mechanisms of identified proteins in lung carcinogenesis.

We linked our findings to translational impacts by characterizing the potential drug targets. From three main drug databases, we identified five putative causal genes linked to nine approved or investigated drugs that may be used for lung cancer treatment. For example, zinc supplementation reduced the viability and increased the apoptotic response in non-small cell lung cancer cell lines (47).

Artenimol is an artemisinin derivative. The anti-malaria compounds artemisinin inhibited the proliferation of A549 and H1299 cells and decreased tumor growth *in vivo* through Wnt/β-catenin inactivation (48).

Moreover, we identified drugs approved for other malignancies that warrant investigation for lung cancer treatment. For example, Zolbetuximab has been approved for treating gastric cancer by the U.S. Food and Drug Administration (FDA). Trifluridine and Tipiracil have received FDA approval for metastatic colorectal and gastric cancer. This repurposing strategy is biologically justified, as a few oncogenic pathways such as KRAS-MAPK, Wnt/β-catenin, and NF-κB are shared between gastric and lung cancer. Future trials and studies are needed to validate the effectiveness and efficiency of these drugs for lung cancer prevention and treatment in real-world settings.

Leveraging multi-layer omics data, we found several gene-protein-cancer trios underlying lung cancer susceptibility. These pathways represent that genetic variants influence lung cancer susceptibility through coordinated mediation in both mRNA expression and protein abundance. Over 60% of proteins with reliable prediction models can be also predicted reliably at their transcriptome levels. The effect sizes between PWAS and TWAS are modestly correlated. However, nearly 40% of proteins showed significant PWAS associations but lacked corresponding TWAS signals at nominal *P* < 0.05, demonstrating that PWAS offers complementary insights. Strikingly, C4B demonstrated opposite effect direction between PWAS and TWAS, which may be attributable to post-transcriptional and post-translational regulatory mechanisms. For example, post-translational modifications influence protein abundance without changing mRNA levels (49). We also found a weak genome-wide protein-mRNA correlation, aligning with findings from the previous studies (9,50), which strongly support that changes in mRNA abundance do not necessarily translate into corresponding changes in protein abundance. These highlighted the necessity of protein-level investigation in order to have a comprehensive understanding of the biological mechanisms of lung cancer susceptibility.

Many proteins detected in lung tissue samples were not detected in plasma. For example, about 20% of our identified proteins using the MS method are not available in plasma proteomics data based on data from the HPA. This means that genetic associations with these proteins could not be discovered by previous blood-based studies, which are mainly based on Olink or SOMAScan platforms. For the proteins that are well-predicted in both blood (51) and lung tissue, we found a moderate correlation in effect size (Spearman’s rho=0.57, *P* <0.001) of associations with lung cancer risk. CTSH, IL16, and TYMP proteins showed significant associations with lung cancer risk in the same directions among both blood and lung tissue. These tissue and blood concordant risk proteins suggest that their blood protein level may serve as potential biologically meaningful noninvasive biomarkers for early detection of lung cancer. Notably, TYMP protein showed a potentially causal association with LUAD risk through our *in silico* analysis.

This significant association of measured protein abundance with lung cancer risk and the concordant effect direction was studied in an external case-control study with 731 smoking-matched participants (52). Combining biological relevance with clinical applicability, this protein is promising for precision medicine and biomarker discovery. Future studies are needed to explore clinical utility and performance in clinical settings.

A major strength of this study is that it is the first large-scale investigation of lung tissue-based protein markers for lung cancer risk by integrating multi-omics data. *In silico* analyses enabled our identification of high-confidence causal proteins. However, several limitations should be noted. First, the lack of normal lung tissue samples from individuals of diverse ancestry restricts our findings to the European ancestry population, which may limit the generalizability of our findings. Second, while we employed internal validation, the absence of an independent external cohort prevents validation of our prediction models and PWAS association results. Third, we only incorporated *cis*-acting genetic variants to construct genetic prediction models for protein abundance. The exclusion of trans-acting variants that regulate protein abundance through protein-protein interactions may underestimate the genetic architecture of protein regulation. Further advanced and computationally efficient methods are needed to leverage the information of *trans*-acting genetic variants in genetic prediction models. Finally, many proteins form protein complexes through protein-protein interaction, thereby impacting lung cancer risk. However, DIA only allows us to detect individual proteins instead of protein complexes. Future research with diverse populations and advanced algorithms is warranted to confirm our findings.

In conclusion, in this first large-scale lung tissue-based PWAS, we identified 29 unique proteins associated with risk for overall lung cancer or its histological subtypes. Further *in silico* analyses prioritized 11 potential causal proteins and identified enrichment in immune-related biological processes and pathways. By leveraging multi-omics data, several gene-protein-cancer trios were uncovered. The putative causal proteins are linked to nine drugs that have been approved or are being investigated in phase III trials. Zinc and Artenimol demonstrated known efficacy, and several gastric cancer treatment drugs may be repurposed for lung cancer treatment. These findings provided valuable insights into lung cancer biology and informed future drug targets and repurposing opportunities.

## Ethics Statement

Deidentified data used in this study were derived from published genome-wide association studies or public data that followed the relevant institutional review boards and participant consent procedures. This study was approved by Vanderbilt University Institutional Review Board.

## Data Availability

The protein prediction models generated in this study will be deposited in Zenodo upon the time of publication. Raw proteomics data from the Vanderbilt Thoracic Biorepository (VTB) used in the present study are available upon a request to the corresponding author, Dr. Qiuyin Cai. The Transdisciplinary Research in Cancer of the Lung team of the International Lung Cancer Consortium (TRICL-ILCCO) GWAS summary statistics were obtained from dbGaP under accession code phs001273.v1.p1. The processed GWAS summary statistics of UK Biobank, FinnGen R12, and the Million Veteran Project were downloaded from https://finngen.gitbook.io/documentation/data-download. Drugs and chemical compounds data were extracted from publicly available databases, including DrugBank (https://go.drugbank.com/), ChEMBL (https://www.ebi.ac.uk/chembl/), and TTD (https://ttd.idrblab.cn/). Blood protein information can be found in The Human Protein Atlas (https://www.proteinatlas.org/). ARIC plasma protein prediction models can be downloaded from http://nilanjanchatterjeelab.org/pwas.

## Code Availability

Publicly available software and packages were used throughout this study according to the developer’s instructions. Adopted parameters were specified in the manuscript. *R version 4.4.0*: https://www.r-project.org/; *R PEER package*: https://github.com/PMBio/peer; *S-PrediXcan*: https://github.com/hakyimlab/MetaXcan; *METAL*: https://github.com/statgen/METAL; *SMR*: https://yanglab.westlake.edu.cn/software/smr/; *R coloc package*: https://github.com/chr1swallace/coloc

## Author Contributions

S.X. J.L. and Q.C. conceived the study and designed the analysis. J. L. and Q.C. supervised the study. S.X., J.S., R.T., D.Y., X.G., W.W., Y.Y., B.L., J.L., and Q.C. analyzed data. S.X., J.S., J.L., and Q.C. interpreted the data and drafted the original manuscript. All the authors significantly contributed to reviewing and revising the manuscript.

## Conflict of Interest

The authors declare no competing interests.

## Funding

This work was supported in part by grant from the National Cancer Institute of the National Institutes of Health (R01CA249863 to Q.C. and J.L.). Data collection and sample preparation were performed by the Survey and Biospecimen Shared Resource which is supported in part by the Vanderbilt-Ingram Cancer Center (P30CA068485).

## Acknowledgement

The authors appreciate the contributions and efforts of the researchers and staff of VTB. We also thank Ms. Kathleen Harmeyer for assistance with manuscript editing and preparation. This work was conducted in part using the resources of the Advanced Computing Center for Research and Education at Vanderbilt University.

## Notes

### Competing Interest Statement

The authors have declared no competing interest.

